# Obesity, Ethnicity, and Covid-19 Mortality: A population-based cohort study of 12.6 Million Adults in England

**DOI:** 10.1101/2021.07.22.21260416

**Authors:** Thomas Yates, Annabel Summerfield, Cameron Razieh, Amitava Banerjee, Yogini Chudasama, Melanie J Davies, Clare Gillies, Nazrul Islam, Claire Lawson, Evgeny Mirkes, Francesco Zaccardi, Kamlesh Khunti, Vahé Nafilyan

**Author notes:** ***Corresponding author*:** Prof Tom Yates; Address: Diabetes Research Centre, University of Leicester, Leicester General Hospital, Leicester, LE5 4PW, UK. joint senior authors (Kamlesh Khunti, Vahé Nafilyan).

## Abstract

**Importance:** Obesity and ethnicity are well characterised risk factors for severe COVID-19 outcomes, but the differential effects of obesity on COVID-19 outcomes by race/ethnicity has not been examined robustly in the general population.

**Objective:** To investigate the association between body mass index (BMI) and COVID-19 mortality across different ethnic groups.

**Design, Setting, and Participants:** This is a retrospective cohort study using linked national Census, electronic health records and mortality data for English adults aged 40 years or older who were alive at the start of pandemic (24^th^ January 2020).

**Exposures:** BMI obtained from electronic health records. Self-reported ethnicity (white, black, South Asian, other) was the effect-modifying variable.

**Main Outcomes and Measures:** COVID-19 related death identified by ICD-10 codes U07.1 or U07.2 mentioned on the death certificate from 24^th^ January 2020 until December 28^th^ 2020.

**Results:** The analysis included white (n = 11,074,708; mean age 61.9 [±13.4] years; 54% women), black (n = 416,542; 56.4 [±11.7] years; 57% women), South Asian (621,691; 55.7 [±12.4] years; 51% women) and other (n = 478,196; 55.3 [±11.6] years; 55% women) ethnicities with linked BMI data. The association between BMI and COVID-19 mortality was stronger in ethnic minority groups. Compared to a BMI of 22.5 kg/m^2^ in white ethnicities, the adjusted HR for COVID-19 mortality at a BMI of 30 kg/m^2^ in white, black, South Asian and other ethnicities was 0.95 (95% CI: 0.87-1.03), 1.72 (1.52-1.94), 2.00 (1.78-2.25) and 1.39 (1.21-1.61), respectively. The estimated risk of COVID-19 mortality at a BMI of 40 kg/m^2^ in white ethnicities (HR = 1.73) was equivalent to the risk observed at a BMI of 30.1 kg/m^2^, 27.0 kg/m^2^, and 32.2 kg/m^2^ in black, South Asian and other ethnic groups, respectively.

**Conclusions:** This population-based study using linked Census and electronic health care records demonstrates that the risk of COVID-19 mortality associated with obesity is greater in ethnic minority groups compared to white populations.

**Question:** Does the association between BMI and COVID-19 mortality vary by ethnicity?

**Findings:** In this study of 12.6 million adults, BMI was associated with COVID-19 in all ethnicities, but with stronger associations in ethnic minority populations such that the risk of COVID-19 mortality for a BMI of 40 kg/m^2^ in white ethnicities was observed at a BMI of 30.1 kg/m^2^, 27.0 kg/m^2^, and 32.2 kg/m^2^ in black, South Asian and other ethnicities, respectively.

**Meaning:** BMI is a stronger risk factor for COVID-19 mortality in ethnic minorities. Obesity management is therefore a priority in these populations.

## INTRODUCTION

Obesity has emerged as one of the most characterised risk factors internationally for coronavirus disease 2019 (COVID-19) severity and mortality in both community and in-patient settings [1-7]. The strong association between obesity and COVID-19 outcomes has been suggested to result from a deleterious change in the role of circulating adipocytokine leading to a pro-inflammatory state with subsequent predisposition to thrombosis, incoordination of innate and adaptive immune responses, inadequate antibody responses, and the cytokine storm [1].

There is growing evidence that the strength of association between BMI and COVID-19 outcomes may be modified by key sociodemographic factors, most notably ethnicity [6,7], which is also an important risk factor of COVID-19 severity and mortality, with risk up to four times greater in black and South Asian ethnicities [8-10]. In a study of 65,932 in-patients admitted with COVID-19 [7], a coding of obesity was associated with a higher risk of intensive care, mechanical ventilation or in-hospital mortality in all ethnic groups, but with the greatest risk observed in black ethnicities with obesity [7]. A community study of 6.9 million adults from general practices in England also found the association between BMI and COVID-19 mortality at the start of pandemic was strongest in black ethnicities [6]. However, whilst ethnicity has been shown to modify associations between BMI and COVID-19 outcomes, previous research has not quantified how this interaction affects both within-ethnicity and between-ethnicity risk across the spectrum of BMI. An early analysis of 5,623 community and in-hospital test results suggested that the risk of SARS-CoV-2 positivity was not different between ethnic groups at low BMI, but was over two fold higher in minority ethnic groups compared to white ethnicities at high BMI [11]. However, this has not been explored in larger representative community cohorts or with COVID-19 outcomes.

Previous analyses with cardiometabolic outcomes have used the differential associations between ethnicity and BMI to calculate thresholds for obesity in minority ethnic groups where risk is equivalent to white ethnicities at established thresholds for obesity (e.g. 30 kg/m^2^) [12-14], with current guidelines suggesting that thresholds for minority ethnic groups should be reduced by 2.5 kg/m^2^ [15,16]. It is unclear whether these guidelines are applicable to COVID-19 outcomes. Therefore elucidating the within and between ethnicity risk with COVID-19 mortality has important implications for public health policy and guidelines in relation to infectious disease.

The aim of this study was to use linked national Census, electronic health care records and mortality datasets to investigate the continuous association between BMI and COVID-19 mortality across different ethnic groups within the general population and to establish equivalency in risk across ethnic groups at established BMI thresholds for class I, II, and III obesity.

## METHODS

### Populations and databases

This analysis uses data from the Office of National Statistics (ONS) Public Health Data Asset, a new linked dataset using the 2011 Census data; all adults within England are required to complete and return Census data by law, with a response rate of 93.9% [17]. The 2011 Census was linked to the General Practice Extraction Service Data for Pandemic Planning and Research (GDPPR) which contains primary care records for all individuals living in England on November 1st 2019, with records being extracted up to December 31st 2019. This dataset was further linked to mortality records and Hospital Episode Statistics (HES). Linkage between clinical and Census datasets was enabled through NHS numbers that are unique to each individual. To obtain NHS numbers for the 2011 Census, the 2011 Census was linked to the 2011-2013 NHS Patient Registers. It was first linked deterministically using 24 different matching keys. Probabilistic matching (Felligi-Sunter method) was then used to match records that were not linked deterministically, using 13 different combinations of personal identifiers [18]. Our analysis was restricted to those over 40 years of age on December 31st 2019 due to poor coverage of BMI values in GDPPR in younger populations.

Of the 32,755,633 people enumerated at the 2011 Census in England and Wales aged ≥40 years on December 31st 2019, 31,498,128 people were linked deterministically or probabilistically to the NHS Patient register, and of these, 27,477,607 individuals were alive on 24^th^ January 2020. As linked family practice data was only available for England, the English population with linked GDPPR data included 24,026,950 people (see sample flow diagram in **Supplementary eFigure 1**). Of these, 12,591,137 (52.4%) had valid BMI data and were included within the primary analysis. The difference in sociodemographic and clinical data between those with and without BMI data are displayed in **Supplementary eTable 2**.

### Exposures

BMI was extracted from GDPPR, reflecting the BMI value coded within primary care that was closest to December 31st 2019. Within the GDPPR extract of primary care records, BMI data were available from January 2010. Participants without a recorded BMI in primary care within this 10 year window were coded as missing. In order to remove outliers and potentially spurious values, a data driven approach was used, restricting the analysis from the 2.5th to 97.5th percentile of the distribution.

### Outcome

COVID-19 related death (either in hospital or out of hospital), defined as confirmed or suspected COVID-19 death, was identified by ICD-10 codes U07.1 or U07.2 mentioned anywhere on the death certificate from 24^th^ January 2020 until December 28^th^ 2020.

### Effect modifier

Self-reported ethnicity was coded from the 2011 Census, which asked respondents to select their ethnicity from 18 categories. For the purposes of this analysis we derived four categories: white (defined as British, Irish, other White), South Asian (Asian/Asian British defined as Indian, Pakistani, Bangladeshi), black (defined as black African, black Caribbean, black British, Other black) or other (all other classifications). Ethnicity was imputed in 3.0% of 2011 Census returns due to item non-response using nearest-neighbour donor imputation, the methodology employed by the Office for National Statistics across all 2011 Census variables [18].

### Covariates

Our analysis included key sociodemographic data including measures of deprivation, household composition, occupation and key worker status, educational attainment and exposure to others, defined within **Supplementary eTable 1**. We also used the linkage to GDPPR and HES to extract data on chronic diseases that have been shown to be associated with COVID-19 outcomes in prediction models [19], see **Supplementary eTable 1**

### Statistical analysis

Cox proportional hazard models were fitted with time to event measured in days from 24^th^ January 2020 to the date of COVID-19 deaths or deaths from other causes or December 28^th^ 2020, whichever came first. Non-COVID-19 mortality was analysed as a censoring event. *A priori* covariates were adjusted for in two models. Model 1 was adjusted for age, sex, ethnicity, geographic region, and other key sociodemographic factors (details provided in **Supplementary eTable 1**). Model 2 was adjusted for the same factors as Model 1, plus included clinical factors (**Supplementary eTable 1**). A BMI by ethnicity interaction term was included in both models. Given the potential for included clinical factors to act as mediators between BMI and COVID-19 mortality, the primary interpretation from the analysis was taken from Model 1. The proportional hazards assumption was assessed visually using log-log survival plots across quartiles of BMI. The strength of interaction was tested using a likelihood-ratio test. Restricted cubic splines were fitted with 3 knots at the 25th (23.2 kg/m^2^), 50th (26.3 kg/m^2^) and 75th (29.8 kg/m^2^) BMI percentiles. A BMI of 22.5 kg/m^2^ (representing a value within the normal range) in white ethnicities as the largest group was specified as the reference to which all other ethnicities and BMI values were compared. Model fit was determined using the concordance statistic (c-index), with values over 0.8 interpreted as a strong model fit.

Values generated by the restricted cubic splines were used to quantify the BMI values in minority ethnicity groups that would produce an equivalent risk to white ethnicities at the thresholds for class I (30 kg/m^2^), II (35 kg/m^2^), and III (40 (kg/m^2^) obesity.

When fitting the Cox models, we included all individuals who died during the analysis period and a weighted random sample of those who did not, with a sampling rate of 1% for those of white British ethnicity and 10% for adults from ethnic minority groups. We applied case weights (defined as the inverse of the sampling rate) to all analyses.

In order to assess the pattern of results across sex and age, analyses were repeated stratified by sex and age (<70 years, ≥ 70 years). In order to assess whether the pattern of results for the broad ethnic categories of white, black, South Asian and other mirrored the pattern of results in more detailed sub-categories, the analysis was repeated using ten categories of ethnicity.

As BMI is likely to be missing not at random and influenced by many factors [20], not all of which were captured in this analysis, multiple imputation of missing data was not attempted. Nevertheless, to assess whether the pattern of missingness varied by ethnic groups, we examined the proportion of missing data by ethnicity across regions. There was no clear systematic pattern of missingness by ethnicity (**supplementary eFigure 2**).

## Results

This analysis included 11,074,708 (53.6% women, 61.9 [±13.4] years) white, 416,542 (57.3% women, 56.4 [±11.7] years) black, 621,691 (51.0% women, 55.7 [±12.4] years) South Asian and 478,196 (54.9% women, 55.3 [±11.6] years) other ethnic groups with linked BMI data. The full descriptive profile of the cohort is displayed in **Table 1**. There were 30,067 (0.27%), 1,208 (0.29%), 1,831 (0.29%), 845 (0.18%) COVID-19 deaths in white, black, South Asian and other ethnic groups, respectively.

**Table 1:** Population characteristics, stratified by ethnicity.

|  |  | **White**<br>**(n = 11,074,708)** | **Black**<br>**(n = 416,542)** | **South Asian**<br>**(n = 621,691)** | **Other**<br>**(n = 478,196)** |
| --- | --- | --- | --- | --- | --- |
| Age (yr) (mean [SD]) |  | 61.9 (13.38) | 56.35 (11.74) | 55.66 (12.37) | 55.25 (11.58) |
| Sex (%) | Male | 5,139,079 (46.40) | 177,981 (42.73) | 304,916 (49.05) | 215,736 (45.11) |
|  | Female | 5,935,629 (53.60) | 238,561 (57.27) | 316,775 (50.95) | 262,460 (54.89) |
| Body Mass Index (kg/m<sup>2</sup>)<br>(mean [SD]) |  | 27.85 (5.69) | 29.34 (5.88) | 27.23 (5.01) | 26.72 (5.2) |
| Region (%) | East | 527,620 (4.76) | 7,958 (1.91) | 16,732 (2.69) | 15,908 (3.33) |
|  | East Midlands | 305,002 (2.75) | 3,706 (0.89) | 9,329 (1.50) | 5,242 (1.10) |
|  | London | 1,404,653 (12.68) | 282,743 (67.88) | 279,403 (44.94) | 240,467 (50.29) |
|  | North East | 477,650 (4.31) | 1,789 (0.43) | 4,940 (0.79) | 6,999 (1.46) |
|  | North West | 2,474,713 (22.35) | 27,346 (6.57) | 94,299 (15.17) | 53,092 (11.10) |
|  | South East | 2,639,897 (23.84) | 34,334 (8.24) | 75,532 (12.15) | 83,377 (17.44) |
|  | South West | 1,015,274 (9.17) | 9,868 (2.37) | 9,654 (1.55) | 16,072 (3.36) |
|  | West Midlands | 1,734,175 (15.66) | 43,509 (10.45) | 124,469 (20.02) | 48,894 (10.22) |
|  | Yorkshire and the Humber | 495,724 (4.48) | 5,289 (1.27) | 7,333 (1.18) | 8,145 (1.70) |
| Urban Rural classification (%) | Rural hamlets and isolated dwellings | 434,588 (3.92) | 899 (0.22) | 1,774 (0.29) | 3,840 (0.80) |
|  | Rural hamlets and isolated dwellings in a sparse setting | 38,604 (0.35) | 29 (0.01) | 36 (0.01) | 206 (0.04) |
|  | Rural town and fringe | 954,956 (8.62) | 2,801 (0.67) | 5,418 (0.87) | 10,626 (2.22) |
|  | Rural town and fringe in a sparse setting | 39,825 (0.36) | 59 (0.01) | 86 (0.01) | 342 (0.07) |
|  | Rural village | 679,768 (6.14) | 1,445 (0.35) | 2,781 (0.45) | 6,134 (1.28) |
|  | Rural village in a sparse setting | 44,749 (0.40) | 36 (0.01) | 50 (0.01) | 258 (0.05) |
|  | Urban city and town | 4,633,331 (41.84) | 54,516 (13.09) | 147,247 (23.68) | 116,791 (24.42) |
|  | Urban city and town in a sparse setting | 23,073 (0.21) | 30 (0.01) | 91 (0.01) | 204 (0.04) |
|  | Urban major conurbation | 4,041,153 (36.49) | 352,179 (84.55) | 458,337 (73.72) | 334,828 (70.02) |
|  | Urban minor conurbation | 184,661 (1.67) | 4,548 (1.09) | 5,871 (0.94) | 4,967 (1.04) |
| Population density (mean [SD]) |  | 3,873.7 (3,929.8) | 9,927.05 (6,316.75) | 8,111.83 (5,966.41) | 7,647.58 (6,045.64) |
| Household deprivation (%) | Not deprived in any dimension | 5,398,007 (48.74) | 150,596 (36.15) | 224,081 (36.04) | 196,694 (41.13) |
|  | Deprived in 1 dimension | 3,407,802 (30.77) | 147,433 (35.39) | 208,303 (33.51) | 158,813 (33.21) |
|  | Deprived in 2 dimensions | 1,629,446 (14.71) | 83,187 (19.97) | 133,715 (21.51) | 84,671 (17.71) |
|  | Deprived in 3 dimensions | 462,202 (4.17) | 27,292 (6.55) | 48,372 (7.78) | 30,195 (6.31) |
|  | Deprived in 4 dimensions | 42,322 (0.38) | 3,585 (0.86) | 5,033 (0.81) | 4,384 (0.92) |
|  | No code | 134,929 (1.22) | 4,449 (1.07) | 2,187 (0.35) | 3,439 (0.72) |
| Material deprivation (IMD decile) (%) | 1 (most deprived) | 825,760 (7.46) | 88,063 (21.14) | 117,028 (18.82) | 58,461 (12.23) |
|  | 2 | 888,014 (8.02) | 103,716 (24.90) | 100,441 (16.16) | 69,802 (14.60) |
|  | 3 | 963,592 (8.70) | 78,278 (18.79) | 79,563 (12.80) | 65,706 (13.74) |
|  | 4 | 1,020,432 (9.21) | 48,339 (11.60) | 68,582 (11.03) | 54,962 (11.49) |
|  | 5 | 1,096,988 (9.91) | 33,198 (7.97) | 61,471 (9.89) | 47,924 (10.02) |
|  | 6 | 1,141,078 (10.30) | 21,532 (5.17) | 53,983 (8.68) | 43,776 (9.15) |
|  | 7 | 1,215,851 (10.98) | 14,015 (3.36) | 39,296 (6.32) | 36,208 (7.57) |
|  | 8 | 1,258,087 (11.36) | 11,301 (2.71) | 34,596 (5.56) | 34,084 (7.13) |
|  | 9 | 1,288,482 (11.63) | 9,923 (2.38) | 32,394 (5.21) | 33,809 (7.07) |
|  | 10 (least deprived) | 1,376,424 (12.43) | 8,177 (1.96) | 34,337 (5.52) | 33,464 (7.00) |
| Approximate social grade (%) | AB | 2,668,369 (24.09) | 71,954 (17.27) | 160,894 (25.88) | 118,489 (24.78) |
|  | C1 | 3,296,637 (29.77) | 131,599 (31.59) | 153,071 (24.62) | 134,981 (28.23) |
|  | C2 | 2,402,096 (21.69) | 76,811 (18.44) | 87,409 (14.06) | 79,408 (16.61) |
|  | D | 2,343,816 (21.16) | 98,784 (23.72) | 182,771 (29.40) | 112,265 (23.48) |
|  | E | 320,788 (2.90) | 34,343 (8.24) | 36,252 (5.83) | 30,694 (6.42) |
|  | No code | 43,002 (0.39) | 3,051 (0.73) | 1,294 (0.21) | 2,359 (0.49) |
| Highest educational attainment (%) | No qualifications | 2,501,606 (22.59) | 66,187 (15.89) | 157,047 (25.26) | 91,730 (19.18) |
|  | 1-4 GCSE/O-levels | 1,523,484 (13.76) | 53,064 (12.74) | 68,560 (11.03) | 45,607 (9.54) |
|  | 5+ GCSE/O-levels | 1,583,286 (14.30) | 55,961 (13.43) | 49,393 (7.94) | 43,763 (9.15) |
|  | Apprenticeship | 467,991 (4.23) | 5,594 (1.34) | 3,795 (0.61) | 4,005 (0.84) |
|  | 2+ A Levels or equivalent | 1,144,859 (10.34) | 39,687 (9.53) | 36,337 (5.84) | 34,826 (7.28) |
|  | Degree or above | 3,338,014 (30.14) | 155,420 (37.31) | 208,981 (33.61) | 179,098 (37.45) |
|  | Other | 515,468 (4.65) | 40,629 (9.75) | 97,578 (15.70) | 79,167 (16.56) |
| Household tenancy (%) | Owned: owned outright | 3,880,716 (35.04) | 42,051 (10.10) | 167,958 (27.02) | 86,111 (18.01) |
|  | Owned: owned with a mortgage or loan | 4,582,819 (41.38) | 129,629 (31.12) | 286,544 (46.09) | 185,759 (38.85) |
|  | Shared ownership | 66,361 (0.60) | 6,149 (1.48) | 2,953 (0.47) | 5,203 (1.09) |
|  | Social rented from council | 699,678 (6.32) | 86,590 (20.79) | 37,233 (5.99) | 45,814 (9.58) |
|  | Other social rented | 686,324 (6.20) | 71,740 (17.22) | 33,825 (5.44) | 40,654 (8.50) |
|  | Private rented | 1,024,833 (9.25) | 72,820 (17.48) | 84,360 (13.57) | 105,581 (22.08) |
|  | Living rent free | 90,975 (0.82) | 4,512 (1.08) | 7,524 (1.21) | 6,715 (1.40) |
|  | No code required | 43,002 (0.39) | 3,051 (0.73) | 1,294 (0.21) | 2,359 (0.49) |
| Type of accommodation (%) | Detached | 3,127,510 (28.24) | 28,945 (6.95) | 113,196 (18.21) | 77,268 (16.16) |
|  | Semi-detached | 3,867,398 (34.92) | 89,608 (21.51) | 195,917 (31.51) | 127,666 (26.70) |
|  | Terraced | 2,601,304 (23.49) | 115,552 (27.74) | 202,687 (32.60) | 125,626 (26.27) |
|  | Flat (purpose built) | 965,900 (8.72) | 148,382 (35.62) | 86,771 (13.96) | 110,458 (23.10) |
|  | Flat (converted) | 277,700 (2.51) | 26,215 (6.29) | 13,453 (2.16) | 24,345 (5.09) |
|  | Flat (commercial building) | 60,018 (0.54) | 3,242 (0.78) | 7,315 (1.18) | 9,006 (1.88) |
|  | Other | 39,949 (0.36) | 149 (0.04) | 165 (0.03) | 388 (0.08) |
|  | No code | 134,929 (1.22) | 4,449 (1.07) | 2,187 (0.35) | 3,439 (0.72) |
| Household size (%) | 1-2 | 7,289,085 (65.82) | 223,481 (53.65) | 176,472 (28.39) | 218,994 (45.80) |
|  | 3-4 | 3,333,717 (30.10) | 151,558 (36.38) | 298,681 (48.04) | 201,504 (42.14) |
|  | 5-6 | 300,440 (2.71) | 33,659 (8.08) | 114,909 (18.48) | 46,748 (9.78) |
|  | 7+ | 16,537 (0.15) | 3,395 (0.82) | 29,442 (4.74) | 7,511 (1.57) |
|  | Missing | 134,929 (1.22) | 4,449 (1.07) | 2,187 (0.35) | 3,439 (0.72) |
| Multigenerational household (%) | No | 9,758,322 (88.11) | 356,325 (85.54) | 452,848 (72.84) | 395,678 (82.74) |
|  | Yes | 1,181,457 (10.67) | 55,768 (13.39) | 166,656 (26.81) | 79,079 (16.54) |
|  | No code | 134,929 (1.22) | 4,449 (1.07) | 2,187 (0.35) | 3,439 (0.72) |
| Household with children (%) | Yes | 1,941,377 (17.53) | 133,773 (32.12) | 262,010 (42.14) | 160,332 (33.53) |
| Overcrowded (%) | Yes | 532,653 (4.81) | 113,070 (27.14) | 128,682 (20.70) | 101,066 (21.13) |
| Key worker type (%) | Education and childcare | 746,622 (6.74) | 19,715 (4.73) | 26,698 (4.29) | 18,542 (3.88) |
|  | Food and necessary goods | 65,869 (0.59) | 1,756 (0.42) | 5,906 (0.95) | 2,678 (0.56) |
|  | Health and social care | 797,565 (7.20) | 64,023 (15.37) | 41,345 (6.65) | 49,803 (10.41) |
|  | Key public services | 185,020 (1.67) | 7,181 (1.72) | 6,891 (1.11) | 6,309 (1.32) |
|  | National and Local Government | 100,392 (0.91) | 4,625 (1.11) | 4,662 (0.75) | 2,792 (0.58) |
|  | Not keyworker | 8,739,732 (78.92) | 307,134 (73.73) | 515,151 (82.86) | 385,328 (80.58) |
|  | Public safety and national security | 158,024 (1.43) | 3,860 (0.93) | 2,879 (0.46) | 3,435 (0.72) |
|  | Transport | 129,584 (1.17) | 4,571 (1.10) | 5,342 (0.86) | 3,444 (0.72) |
|  | Utilities and communication | 151,900 (1.37) | 3,677 (0.88) | 12,817 (2.06) | 5,865 (1.23) |
| Key worker in household (%) | No code | 134,929 (1.22) | 4,449 (1.07) | 2,187 (0.35) | 3,439 (0.72) |
|  | No | 7,170,389 (64.75) | 247,815 (59.49) | 405,434 (65.21) | 318,454 (66.59) |
|  | Yes | 3,769,390 (34.04) | 164,278 (39.44) | 214,070 (34.43) | 156,303 (32.69) |
| Proximity to others* (mean [SD]) |  | 58.41 (18.86) | 58.95 (25.13) | 51.53 (28.1) | 56.14 (25.95) |
| Exposure to disease* (mean [SD]) |  | 18.98 (20.68) | 26.7 (26.67) | 17.3 (22.35) | 21.3 (24.65) |
| Chronic Kidney disease (%) | None, CKD 1-2 | 10,775,193 (97.30) | 405,526 (97.36) | 607,621 (97.74) | 471,354 (98.57) |
|  | CKD 3 | 262,284 (2.37) | 8,764 (2.10) | 11,140 (1.79) | 5,586 (1.17) |
|  | CKD 4 | 26,660 (0.24) | 1,237 (0.30) | 1,674 (0.27) | 659 (0.14) |
|  | CKD 5 | 10,571 (0.10) | 1,015 (0.24) | 1,256 (0.20) | 597 (0.12) |
| Learning disability (%) | None | 10,935,617 (98.74) | 412,954 (99.14) | 616,307 (99.13) | 474,373 (99.20) |
|  | Learning disability | 135,253 (1.22) | 3,478 (0.83) | 5,244 (0.84) | 3,703 (0.77) |
|  | Down's syndrome | 3,838 (0.03) | 110 (0.03) | 140 (0.02) | 120 (0.03) |
| Cancer and immunosuppression (%) | Blood cancer | 152,888 (1.38) | 3,907 (0.94) | 5,236 (0.84) | 4,033 (0.84) |
|  | Respiratory cancer | 4,666 (0.04) | 239 (0.06) | 146 (0.02) | 112 (0.02) |
|  | Taking immunosuppressants | 3,244 (0.03) | 62 (0.01) | 133 (0.02) | 62 (0.01) |
|  | Taking anti-leukotriene or long acting beta2-agonists | 1,006,065 (9.08) | 21,129 (5.07) | 47,665 (7.67) | 29,058 (6.08) |
|  | Taking oral steroids in the last 6 months | 188,513 (1.70) | 3,331 (0.80) | 6,555 (1.05) | 3,724 (0.78) |
| Other comorbidities (%) | Cerebral Palsy | 1,738 (0.02) | 53 (0.01) | 102 (0.02) | 46 (0.01) |
|  | Asthma | 1,355,348 (12.24) | 43,649 (10.48) | 80,742 (12.99) | 54,625 (11.42) |
|  | Atrial Fibrillation | 567,932 (5.13) | 5,778 (1.39) | 8,641 (1.39) | 7,682 (1.61) |
|  | Coronary heart disease | 756,198 (6.83) | 11,536 (2.77) | 50,263 (8.08) | 21,978 (4.60) |
|  | COPD | 544,244 (4.91) | 5,060 (1.21) | 9,860 (1.59) | 8,026 (1.68) |
|  | Cystic fibrosis or bronchiectasis or alveolitis | 179,952 (1.62) | 4,532 (1.09) | 6,336 (1.02) | 4,350 (0.91) |
|  | Dementia | 204,040 (1.84) | 5,910 (1.42) | 6,231 (1.00) | 3,860 (0.81) |
|  | Diabetes | 1,245,180 (11.24) | 82,220 (19.74) | 174,601 (28.08) | 82,897 (17.34) |
|  | Epilepsy | 138,698 (1.25) | 3,674 (0.88) | 4,917 (0.79) | 3,831 (0.80) |
|  | Heart failure | 267,535 (2.42) | 6,447 (1.55) | 10,927 (1.76) | 5,251 (1.10) |
|  | Liver cirrhosis | 39,354 (0.36) | 915 (0.22) | 1,832 (0.29) | 1,414 (0.30) |
|  | Neurological disease | 12,318 (0.11) | 368 (0.09) | 535 (0.09) | 380 (0.08) |
|  | Parkinson's disease | 57,315 (0.52) | 1,076 (0.26) | 2,163 (0.35) | 1,339 (0.28) |
|  | Peripheral vascular disease | 155,578 (1.40) | 2,697 (0.65) | 4,149 (0.67) | 2,379 (0.50) |
|  | Fracture of hip, wrist, spine or humerus | 17,341 (0.16) | 81 (0.02) | 406 (0.07) | 203 (0.04) |
|  | Rheumatoid arthritis or SLE | 146,556 (1.32) | 4,244 (1.02) | 9,363 (1.51) | 4,949 (1.03) |
|  | Severe mental illness | 2,202,390 (19.89) | 56,330 (13.52) | 80,841 (13.00) | 73,540 (15.38) |
|  | Solid organ transplant | 1,251 (0.01) | 105 (0.03) | 115 (0.02) | 56 (0.01) |
|  | Stroke or TIA | 445,678 (4.02) | 11,534 (2.77) | 16,798 (2.70) | 9,285 (1.94) |
|  | Thrombosis or pulmonary embolus | 2,904 (0.03) | 27 (0.01) | 179 (0.03) | 62 (0.01) |

Data as number (column %) or mean (SD).
\* = score from 0 (no exposure) to 100 (maximum exposure).

BMI was associated with COVID-19 mortality in all ethnic groups (**Figure 1**). However, compared to white ethnicities, the dose-response association was more pronounced in black, South Asian and other ethnic groups (P < 0.001 for interaction). At a BMI of 22.5 kg/m^2^, the risk of COVID-19 mortality was moderately elevated in black (HR: 1.19; 95% CI 1.01, 1.41), South Asian (HR: 1.44; 1.26, 1.64), and other (HR: 1.17; 1.00, 1.38) ethnicities compared to white ethnicities, after adjusting for key sociodemographic factors. However, the difference in risk between ethnic groups was magnified at higher BMI. Compared to the reference of BMI of 22.5 kg/m^2^ in white ethnicities, the HR for COVID-19 mortality at a BMI of 30 kg/m^2^ in white, black, South Asian and other ethnicities was 0.95 (0.87, 1.03), 1.72 (1.52, 1.94), 2.00 (1.78, 2.25) and 1.39 (1.21, 1.61), respectively. For a BMI of 35 kg/m^2^ the HRs for white, black, South Asian and other ethnicities were 1.24 (1.34, 1.14), 2.38 (2.72, 2.08), 3.02 (2.64,3.45) and 2.29 (1.93, 2.72), respectively. For a BMI of 40 kg/m^2^ the HRs for white, black, South Asian and other ethnicities were 1.73 (1.59, 1.91), 3.01 (2.32, 3.90), 5.25 (4.06, 6.79) and 3.89 (2.72, 5.54), respectively.

**Figure 1.**
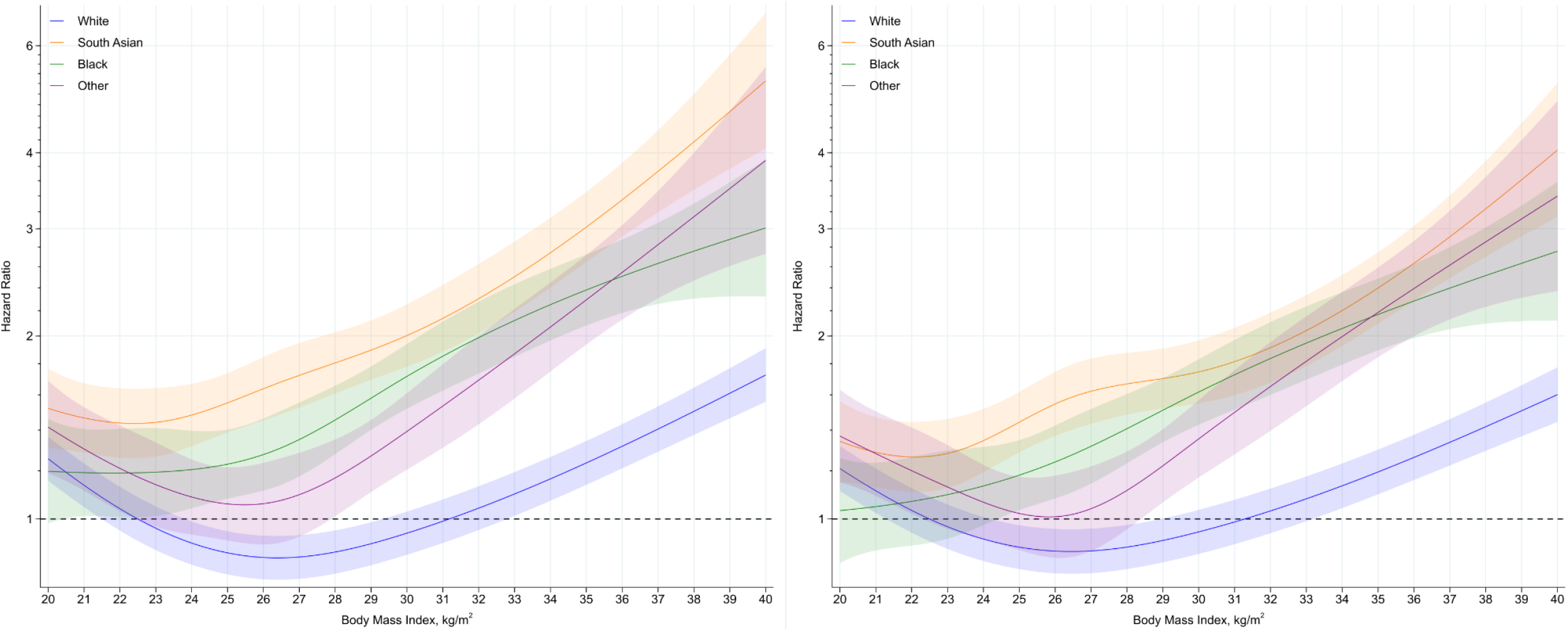
Association of BMI with COVID-19 mortality by ethnicity. Panel A (left) adjusted for: age, sex and demographic variables [detailed in Supplementary eTable 1]. The c-index for the model was 0.902, indicating a strong fit. Panel B (right) adjusted for: age, sex, demographic variables and health and disease status [detailed in Supplementary eTable 1]. The c-index for the model fit was 0.920, indicating a strong fit. Reference (HR = 1) were placed at BMI of 22.5 kg/m^2^ for white ethnicties. HR (lines) and confidence intervals (CI; areas) are plotted across continuous BMI values (x axes) between the 2.5th (20 kg/m^2^) and 97.5th (40 kg/m^2^) centile of the distribution. Shaded area as 95% CI.

The equivalent risk to white ethnicities at a BMI of 35 kg/m^2^ (HR = 1.24) was observed at BMI values of 25.2 (21.5, 27.6) kg/m^2^ and 28.7 (26.0, 30.3) kg/m^2^ in black and other ethnicities, respectively (**Figure 2**); risk equivalence was not possible for South Asian ethnicities where COVID-19 mortality risk was greater than this value even at low BMI. The equivalent risk of COVID-19 mortality in white ethnicities at a BMI of 40 kg/m^2^ (HR = 1.73) was observed at a BMI of 30.1 (28.6, 31.9) kg/m^2^, 27.0 (24.9, 29.4) kg/m^2^, and 32.2 (30.6, 33.9) kg/m^2^ in black, South Asian and other ethnicities, respectively (**Figure 2**). All ethnic minority groups at any BMI value had a higher risk of COVID-19 mortality than white ethnicities at a BMI of 30 kg/m^2^.

**Figure 2.**
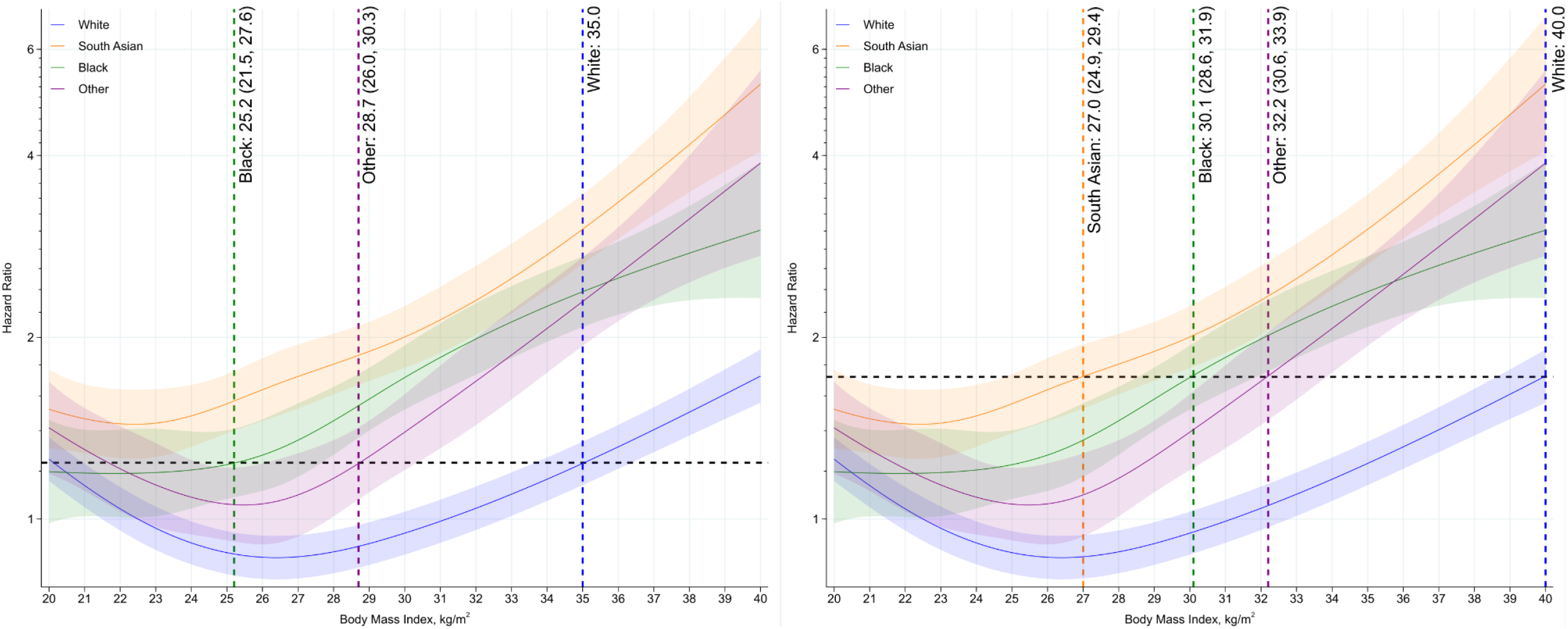
BMI equivalency values for Class II and Class III obesity thresholds across ethnic groups. Panel A (left): BMI values in ethnic minority groups for equivalent risk to white ethnicities at a BMI of 35 kg/m^2^ [HR = 1.24 (1.34, 1.14)]. Model adjusted for: age, sex and demographic variables [detailed in Supplementary eTable 1]. Panel B (right): BMI values in ethnic minority groups for equivalent risk to white ethnicities at a BMI of 40 kg/m^2^ [HR = 1.73 (1.59, 1.91)]. Model adjusted for: age, sex and demographic variables [detailed in Supplementary eTable 1].

The results were similar after further adjustment for clinical factors (model 2 shown in **Figure 1**). With this further adjustment, an equivalent risk to white ethnicities at a BMI of 40 kg/m^2^ was observed at BMI values of 29.9 (28.0, 32.0) kg/m^2^, 26.7 (24.9, 31.1) kg/m^2^, and 31.7 (30.2, 33.5) kg/m^2^ in black, South Asian and other ethnicities references, respectively.

Associations between BMI and COVID-19 mortality were consistent across men and women (**supplementary eFigure 3-4**), but there was evidence that the association was stronger and differences between ethnic groups were more pronounced in those under 70 years of age. In this age group, the risk of COVID-19 mortality at a BMI of 40 kg/m^2^ increased to 2.71 (2.17, 3.38), 6.99 (4.78, 10.22), 9.48 (6.53, 13.75) and 7.95 (5.05, 12.51) in white, black, South Asian and other ethnicities, respectively (model 1), compared to the estimated risk at a BMI of 22.5 kg/m^2^ in white ethnicities (**supplementary eFigure 5-6**).

When analysis was repeated using a broader range of ethnic classification, the pattern of results mirrored the main finding with South Asian ethnicities (Bangladeshi and Pakistani) having the greatest risk at higher BMI **supplementary eFigure 7**. White ethnicities had the lowest risk. However, Chinese ethnicities may not reflect the wider trend for other ethnic minority groups, with associations similar to white ethnicities.

## DISCUSSION

In 12.6 million adults with linked Census, electronic health care records and mortality data, BMI was associated with COVID-19 mortality amongst all ethnic groups with a stronger association in minority ethnic groups. Compared to the reference of a BMI of 22.5 kg/m^2^ in white ethnicities, the risk of COVID-19 mortality at a BMI of 40 kg/m^2^ was 1.73 times higher in white ethnicities, but between 3.01 to 5.25 times higher in ethnic minority populations, with the greatest risk observed in people of South Asian ethnicity. At a BMI of 40.0 kg/m^2^ in white ethnicities, equivalent risk of COVID-19 mortality was observed at BMI thresholds of 30.1, 27.0 and 32.2 kg/m^2^ in black, South Asian and other ethnicities. All ethnic minority groups, even at low BMI, had a higher risk of COVID-19 mortality than white ethnicities at a BMI of 30 kg/m^2^.

To our knowledge, this is the first large-scale population-based study to show the continuous association between BMI and COVID-19 mortality across different ethnic groups on a population level, and to provide BMI values that provide equivalent risk at commonly used thresholds for obesity classifications. Our findings are consistent with previous observations in a community setting at the start of the pandemic and a later in-hospital study [6,7], which also observed an interaction between BMI and ethnicity with COVID-19 outcomes. We extend these previous studies by quantifying the shape of the interaction across a continuous measure of BMI using linked Census and health care records up to the end of 2020, which allowed us to adjust for detailed sociodemographic characteristics and comorbidities in a population-level dataset. Our findings suggest that, unlike other health outcomes such as type 2 diabetes [12-14], it may not be possible to achieve BMI threshold equivalency in the risk of COVID-19 mortality for class I obesity. Current guidelines suggest that BMI threshold for obesity classifications should be reduced by 2.5 kg/m^2^ in minority ethnic groups [15,16]. This study suggests that applying these criteria to COVID-19 mortality will only have a marginal impact and still produce thresholds where risk is substantially elevated in minority ethnic communities compared to white ethnicities.

Our finding that the association between BMI and COVID-19 mortality was stronger in those under 70 years of age is consistent with previous observations from the United States and Europe and provides further evidence for the importance of BMI as a risk factor in younger populations [5-7], especially those from minority ethnic groups where the risk with obesity was greatest.

The reasons underpinning the observed ethnicity by obesity interaction are unclear. It has previously been suggested that ethnic minorities may have a stronger innate inflammatory response to viral infection or chronic disease [21-23], thus potentially increasing the risk of severe COVID-19 [21]. It is possible that the presence of greater levels of adiposity interacts with and accelerates this inflammatory response in ethnic minorities [7]. However, unlike previous findings from the start of the pandemic or from hospital settings where the risk with obesity was found to be greatest in black ethnicities [6,7], this research using national level data from primary care during the first year of the pandemic suggests that the risk with obesity is greater in all minority ethnic groups compared to white ethnicities, with South Asian ethnicities at greatest risk. This mirrors the interaction between ethnicity and BMI with cardiometabolic disease where risk has also consistently been found to be greatest in South Asian ethnicities [12-14]. Further research in this area, including the potential of genetic and epigenetic factors, is warranted.

A major strength is the large population level dataset linking national Census and health care record data making it the largest analysis of its kind to date. Linkage between clinical records and Census data allowed for the extraction of BMI from primary care records and ethnicity from Census data, which is a major strength as ethnicity is not universally coded within primary care [24], with a previous study investigating COVID-19 risk factors in England finding ethnicity coding was missing in over 25% of clinical records [10]. We were also able to extract detailed descriptive and covariate data from a wide range of sociodemographic and clinical factors, allowing for the adjustment of potentially confounding variables including household and area indicators of deprivation and established clinical risk factors for COVID-19 mortality. However, there were limitations. Most notably, this analysis is generalisable to the 52.4% of the English population with coded BMI data within their health care records in the 10 years preceding the pandemic. In England, height and weight are collected as part of routine care by trained staff using medical grade equipment. Nevertheless, family practice incentivisation schemes and differential take-up rates to population level vascular screening programmes means that data is not missing at random [20]. Previous analysis has shown that women, those who attend their family doctor more often, who come from more deprived areas, who have a high or low BMI and have a greater number of comorbidities are more likely to have a coded BMI value [20]. Nevertheless, it has been demonstrated that missing data within clinical records can provide unbiased estimates of adjusted exposure-outcome associations under a wide range of missing data assumptions [25]. In addition, primary care data in England provide some of the most detailed electronic health-care records internationally and are routinely used to identify individuals at risk of chronic and infectious diseases, including COVID-19 mortality [19, 26], giving this study real world utility. This study utilized data from the 2011 Census, therefore any sociodemographic changes within the last decade will not be reflected in the analysis. Although we adjusted for factors related to the risk of SARS-CoV-2 exposure, including household composition, key worker status and exposure to others, it is not possible to verify whether the associations observed with BMI and ethnicity were due to greater disease severity, greater SARS-CoV-2 exposure and infection rates, or a combination of both. Therefore results should be interpreted simply as the population level risk of dying from COVID-19 during the first year of pandemic.

In conclusion, this study of linked Census, electronic health records and mortality data demonstrated a notable interaction between ethnicity and obesity in the risk of COVID-19 mortality, with obesity having a stronger association in all ethnic minorities compared to white ethnicities. The risk of COVID-19 mortality was higher in all minority ethnic groups, even with low BMI, compared to white ethnicities at a BMI of 30 kg/m^2^, suggesting that risk equivalence at the threshold for class I obesity is not possible. These results further emphasise the importance of public health messages to reduce levels of obesity within the population, particularly within ethnic minorities. Future work is needed to investigate how these risk factors interact with post COVID-19 vaccination infection and mortality risk.

## Data Availability

Analysed data is controlled by the Office of National Statistics, UK. Data fields and access request forms can be viewed centrally though the HDR-UK Innovation Gateway https://www.healthdatagateway.org/

## Author contributions

TY, KK, CR, VN developed the research question. TY, CR, VN developed the statistical analysis plan. AS undertook the statistical analysis with support from VN; both had access to the data. TY drafted the manuscript. All authors contributed to the research design and revised the manuscript for important intellectual content.

