## Supplementary data for "Obesity, Ethnicity, and Covid-19 Mortality: A population-based cohort study of 12.6 Million Adults in England"

### Supplementary Material

**eTable 1: Included covariates.**

| Variable | Coding | Model included |
|---|---|---|
| **Geographical variables** | | |
| Region | Dummy variables representing region of residence within England (South East, London, North West, East of England, West Midlands, South West, Yorkshire and the Humber, East Midlands, North East) | 1,2 |
| Population density of Lower Super Output Area (see table note) | Second-order polynomial, allowing for a different slope beyond the 99th percentile of the distribution to account for extreme values | 1,2 |
| Rural urban classification | Rural hamlets and isolated dwellings, Rural hamlets and isolated dwellings in a sparse setting, Rural town and fringe, Rural town and fringe in a sparse setting, Rural village, Rural village in a sparse setting, Urban city and town, Urban city and town in a sparse setting, Urban major conurbation, Urban minor conurbation | 1,2 |
| **Socio-economic variables** | | |
| Index of Multiple Deprivation (IMD)) | Dummy variables representing deciles of deprivation – from 1 (most deprived) to 10 (least deprived) | 1,2 |
| Household deprivation (see table note) | Not deprived, deprived in one dimension, deprived in two dimensions, deprived in three dimensions, deprived in four dimensions | 1,2 |
| Household tenure | Own outright, own with mortgage, social rented, private rented, other | 1,2 |
| Approximate Social Grade of the household reference person (see table note) | AB Higher and intermediate managerial/administrative/professional; C1 Supervisory, clerical, junior managerial/administrative/professional; C2 Skilled manual workers; D Semi-skilled and unskilled manual workers; E On state benefit, unemployed, lowest grade workers (Based on household tenure for people aged 75 or over) | 1,2 |

| | | |
|---|---|---|
| Level of highest qualification | Degree, A-level or equivalent, GCSE or equivalent, no qualification | 1,2 |
| **Household variables** | | |
| Household size | 1-2 people, 3-4 people, 5-6 people, 7+ people | 1,2 |
| Multigenerational household | Dummy for households with at least one person 65+ and someone at least 20 years younger | 1,2 |
| Household with children | At least one child aged 9 to 18 | 1,2 |
| **Occupational exposure variables** (see table note) | | |
| Key worker type | Education & childcare, food & necessity goods, health & social care, public services, national & local government, public safety & national security, transport, utilities & communication, not a key worker | 1,2 |
| Key worker in the household | Yes, no | 1,2 |
| Exposure to disease | Score ranging from 0 (no exposure) to 100 (maximum exposure), derived from O*NET data [12] | 1,2 |
| Proximity to others | Score ranging from 0 (no exposure) to 100 (maximum exposure), derived from O*NET data [12] | 1,2 |
| Household exposure to disease | Maximum 'exposure to disease' score within each household | 1,2 |
| Household proximity to others | Maximum of 'proximity to others' score within each household | 1,2 |
| **Health-related variables** | | |

| | | |
|---|---|---|
| Chronic kidney disease (CKD) | No CKD, CKD3, CKD4, CKD5 | 2 |
| Learning disability | No learning disability, Down's Syndrome, other learning disability | 2 |
| | | 2 |
| Cancer and immunosuppression | Dummies for blood cancer, solid organ transplant , Prescribed immunosuppressant medication by GP , Prescribed leukotriene or long-acting beta blockers, Prescribed regular prednisolone , | 2 |
| Other conditions | Diabetes, Chronic obstructive pulmonary disease (COPD), Asthma , Rare pulmonary diseases , Pulmonary hypertension or pulmonary fibrosis , Coronary heart disease , Stroke , Atrial Fibrillation , Congestive cardiac failure , Venous thromboembolism , Peripheral vascular disease , Congenital heart disease , Dementia , Parkinson's disease , Epilepsy , Rare neurological conditions , Cerebral palsy , Severe mental illness (bipolar disorder, schizophrenia, severe depression), Osteoporotic fracture , Rheumatoid arthritis or Systemic lupus erythematosus , Cirrhosis of the liver | 2 |

Table information: There are 32,844 Lower Super Output Area (LSOA) areas in England, with a mean population of 1500 and a minimum of 1000. We calculated density as LSOA population divided by LSOA area. Household deprivation is defined across four dimensions: employment (at least one household member is unemployed or with long-term sickness, not including full-time students); education (no household member has at least Level 2 education, and no one aged 16-18 years is a full-time student); health and disability (at least one household member reported their health staus as being 'bad'/'very bad' or has a long-term health problem); and housing (the household's accommodation is overcrowded, with an occupancy rating -1 or less, or is in a shared dwelling, or has no central heating). Approximate Social Grade is a socio-economic classification based on the occupation, employment, qualification, and tenure of the household reference person. Key worker type is defined based on the occupation and industry code. 'Exposure to disease' and 'proximity to others' are derived from the O*NET database [1], which collects a range of information about individual working conditions and the day-to-day employment roles. To calculate the proximity and exposure measures, the questions asked were: i) How physically close to other people are you when you perform your current job? ii) How often does your current job require that you be exposed to diseases or infection? Scores ranging from 0 (no exposure) to 100 (maximum exposure) were calculated based on these questions [1]

1. Office for National Statistics. Which occupations have the highest potential exposure to the coronavirus (COVID-19)? 2020. https://www.ons.gov.uk/employmentandlabourmarket/peopleinwork/employmentandempl

oyeetypes/articles/whichoccupationshavethehighestpotentialexposuretothecoronaviruscovid19/2020-05-11 (11 August 2020, date last accessed)

**eTable 2. Comparison of demographic and medical characteristics by those with and without valid body mass index data.**

| Variable | Value | Cases without BMI | Cases with BMI |
|---|---|---|---|
| Ethnicity (%) | Black | 209,977 (1.84) | 416,542 (3.31) |
| | Other | 319,432 (2.79) | 478,196 (3.80) |
| | South Asian | 442,014 (3.87) | 621,691 (4.94) |
| | White | 10,464,547 (91.51) | 11,074,708 (87.96) |
| Age (yr) (mean [SD]) | | 60.85 (13.33) | 61.16 (13.37) |
| Sex (%) | Male | 5,506,657 (48.15) | 5,837,712 (46.36) |
| | Female | 5,929,313 (51.85) | 6,753,425 (53.64) |
| Region (%) | East | 2,245,903 (19.64) | 568,218 (4.51) |
| | East Midlands | 1,856,050 (16.23) | 323,279 (2.57) |
| | London | 714,638 (6.25) | 2,207,266 (17.53) |
| | North East | 731,420 (6.40) | 491,378 (3.90) |
| | North West | 533,380 (4.66) | 2,649,450 (21.04) |
| | South East | 1,284,037 (11.23) | 2,833,140 (22.50) |
| | South West | 1,581,588 (13.83) | 1,050,868 (8.35) |
| | West Midlands | 564,979 (4.94) | 1,951,047 (15.50) |
| | Yorkshire and the Humber | 1,923,975 (16.82) | 516,491 (4.10) |
| Urban Rural classification (%) | Rural hamlets and isolated dwellings | 415,515 (3.63) | 441,101 (3.50) |
| | Rural hamlets and isolated dwellings in a sparse setting | 38,287 (0.33) | 38,875 (0.31) |
| | Rural town and fringe | 1,280,199 (11.19) | 973,801 (7.73) |
| | Rural town and fringe in a sparse setting | 55,457 (0.48) | 40,312 (0.32) |
| | Rural village | 783,065 (6.85) | 690,128 (5.48) |
| | Rural village in a sparse setting | 42,543 (0.37) | 45,093 (0.36) |
| | Urban city and town | 5,594,085 (48.92) | 4,951,885 (39.33) |
| | Urban city and town in a sparse setting | 21,287 (0.19) | 23,398 (0.19) |
| | Urban major conurbation | 2,551,576 (22.31) | 5,186,497 (41.19) |
| | Urban minor conurbation | 653,956 (5.72) | 200,047 (1.59) |
| Population density (mean [SD]) | | 3,668.58 (3,936.75) | 4,426.54 (4,514.81) |
| Household deprivation (%) | Not deprived in any dimension | 5,485,531 (47.97) | 5,969,378 (47.41) |
| | Deprived in 1 dimension | 3,574,785 (31.26) | 3,922,351 (31.15) |
| | Deprived in 2 dimensions | 1,714,038 (14.99) | 1,931,019 (15.34) |
| | Deprived in 3 dimensions | 482,235 (4.22) | 568,061 (4.51) |
| | Deprived in 4 dimensions | 44,150 (0.39) | 55,324 (0.44) |

| | | | |
|---|---|---|---|
| | No code | 135,231 (1.18) | 145,004 (1.15) |
| IMD decile (%) | 1 (most deprived) | 862,777 (7.54) | 1,089,312 (8.65) |
| | 2 | 902,810 (7.89) | 1,161,973 (9.23) |
| | 3 | 1,008,728 (8.82) | 1,187,139 (9.43) |
| | 4 | 1,112,875 (9.73) | 1,192,315 (9.47) |
| | 5 | 1,188,222 (10.39) | 1,239,581 (9.84) |
| | 6 | 1,252,507 (10.95) | 1,260,369 (10.01) |
| | 7 | 1,290,824 (11.29) | 1,305,370 (10.37) |
| | 8 | 1,296,152 (11.33) | 1,338,068 (10.63) |
| | 9 | 1,299,601 (11.36) | 1,364,608 (10.84) |
| | 10 (least deprived) | 1,221,474 (10.68) | 1,452,402 (11.54) |
| Approximate social grade (%) | AB | 2,551,383 (22.31) | 3,019,706 (23.98) |
| | C1 | 3,308,971 (28.93) | 3,716,288 (29.52) |
| | C2 | 2,571,883 (22.49) | 2,645,724 (21.01) |
| | D | 2,620,394 (22.91) | 2,737,636 (21.74) |
| | E | 340,823 (2.98) | 422,077 (3.35) |
| | No code | 42,516 (0.37) | 49,706 (0.39) |
| Highest educational attainment (%) | No qualifications | 2,598,176 (22.72) | 2,816,570 (22.37) |
| | 1-4 GCSE/O-levels | 1,632,648 (14.28) | 1,690,715 (13.43) |
| | 5+ GCSE/O-levels | 1,647,767 (14.41) | 1,732,403 (13.76) |
| | Apprenticeship | 487,068 (4.26) | 481,385 (3.82) |
| | 2+ A Levels or equivalent | 1,179,788 (10.32) | 1,255,709 (9.97) |
| | Degree or above | 3,262,677 (28.53) | 3,881,513 (30.83) |
| | Other | 627,846 (5.49) | 732,842 (5.82) |
| Household tenancy (%) | Owned: owned outright | 3,805,386 (33.28) | 4,176,836 (33.17) |
| | Owned: owned with a mortgage or loan | 4,813,630 (42.09) | 5,184,751 (41.18) |
| | Shared ownership | 69,778 (0.61) | 80,666 (0.64) |
| | Social rented from council | 817,907 (7.15) | 869,315 (6.90) |
| | Other social rented | 618,705 (5.41) | 832,543 (6.61) |
| | Private rented | 1,165,317 (10.19) | 1,287,594 (10.23) |
| | Living rent free | 102,731 (0.90) | 109,726 (0.87) |
| | No code required | 42,516 (0.37) | 49,706 (0.39) |
| Type of accommodation (%) | Detached | 3,451,424 (30.18) | 3,346,919 (26.58) |
| | Semi-detached | 3,936,483 (34.42) | 4,280,589 (34.00) |

| | | | |
|---|---|---|---|
| | Terraced | 2,647,830 (23.15) | 3,045,169 (24.19) |
| | Flat (purpose built) | 927,016 (8.11) | 1,311,511 (10.42) |
| | Flat (converted) | 234,296 (2.05) | 341,713 (2.71) |
| | Flat (commercial building) | 66,691 (0.58) | 79,581 (0.63) |
| | Other | 36,999 (0.32) | 40,651 (0.32) |
| | No code | 135,231 (1.18) | 145,004 (1.15) |
| Household size (%) | 1-2 | 7,277,431 (63.64) | 7,908,032 (62.81) |
| | 3-4 | 3,576,237 (31.27) | 3,985,460 (31.65) |
| | 5-6 | 403,703 (3.53) | 495,756 (3.94) |
| | 7+ | 43,368 (0.38) | 56,885 (0.45) |
| | Missing | 135,231 (1.18) | 145,004 (1.15) |
| Multigenerational household (%) | No | 10,054,231 (87.92) | 10,963,173 (87.07) |
| | Yes | 1,246,508 (10.90) | 1,482,960 (11.78) |
| | No code | 135,231 (1.18) | 145,004 (1.15) |
| Household with children (%) | Yes | 2,232,421 (19.52) | 2,497,492 (19.84) |
| Overcrowded (%) | Yes | 648,996 (5.68) | 875,471 (6.95) |
| Key worker type (%) | Education and childcare | 708,038 (6.19) | 811,577 (6.45) |
| | Food and necessary goods | 92,675 (0.81) | 76,209 (0.61) |
| | Health and social care | 856,170 (7.49) | 952,736 (7.57) |
| | Key public services | 173,130 (1.51) | 205,401 (1.63) |
| | National and Local Government | 93,822 (0.82) | 112,471 (0.89) |
| | Not keyworker | 9,061,425 (79.24) | 9,947,345 (79.00) |
| | Public safety and national security | 163,166 (1.43) | 168,198 (1.34) |
| | Transport | 139,617 (1.22) | 142,941 (1.14) |
| | Utilities and communication | 147,927 (1.29) | 174,259 (1.38) |
| Key worker in household (%) | No code | 135,231 (1.18) | 145,004 (1.15) |
| | No | 7,411,170 (64.81) | 8,142,092 (64.67) |
| | Yes | 3,889,569 (34.01) | 4,304,041 (34.18) |
| Proximity to others* (mean [SD]) | | 58.49 (19.41) | 58.01 (20.02) |
| Exposure to disease* (mean [SD]) | | 19.16 (21.12) | 19.24 (21.2) |
| Chronic Kidney disease (%) | None, CKD 1-2 | 11,267,353 (98.53) | 12,259,694 (97.37) |
| | CKD 3 | 146,007 (1.28) | 287,774 (2.29) |
| | CKD 4 | 14,677 (0.13) | 30,230 (0.24) |

| | | | |
|---|---|---|---|
| | CKD 5 | 7,933 (0.07) | 13,439 (0.11) |
| Learning disability (%) | No | 11,307,924 (98.88) | 12,439,251 (98.79) |
| | Learning disability | 125,410 (1.10) | 147,678 (1.17) |
| | Down's syndrome | 2,636 (0.02) | 4,208 (0.03) |
| Cancer and immunosuppression (%) | Blood cancer | 145,712 (1.27) | 166,064 (1.32) |
| | Respiratory cancer | 4,309 (0.04) | 5,163 (0.04) |
| | Taking immunosuppressants | 3,377 (0.03) | 3,501 (0.03) |
| | Taking anti-leukotriene or long acting beta2-agonists | 941,990 (8.24) | 1,103,917 (8.77) |
| | Taking oral steroids in the last 6 months | 179,500 (1.57) | 202,123 (1.61) |
| Other comorbidities (%) | Cerebral Palsy | 1,574 (0.01) | 1,939 (0.02) |
| | Asthma | 1,227,304 (10.73) | 1,534,364 (12.19) |
| | Atrial Fibrillation | 497,090 (4.35) | 590,033 (4.69) |
| | Coronary heart disease | 703,165 (6.15) | 839,975 (6.67) |
| | COPD | 479,764 (4.20) | 567,190 (4.50) |
| | Cystic fibrosis or bronchiectasis or alveolitis | 155,800 (1.36) | 195,170 (1.55) |
| | Dementia | 194,006 (1.70) | 220,041 (1.75) |
| | Diabetes | 1,268,015 (11.09) | 1,584,898 (12.59) |
| | Epilepsy | 121,092 (1.06) | 151,120 (1.20) |
| | Heart failure | 248,579 (2.17) | 290,160 (2.30) |
| | Liver cirrhosis | 35,152 (0.31) | 43,515 (0.35) |
| | Neurological disease | 11,339 (0.10) | 13,601 (0.11) |
| | Parkinson's disease | 51,350 (0.45) | 61,893 (0.49) |
| | Peripheral vascular disease | 137,143 (1.20) | 164,803 (1.31) |
| | Fracture of hip, wrist, spine or humerus | 10,999 (0.10) | 18,031 (0.14) |
| | Rheumatoid arthritis or SLE | 133,379 (1.17) | 165,112 (1.31) |
| | Severe mental illness | 2,220,379 (19.42) | 2,413,101 (19.17) |
| | Solid organ transplant | 1,296 (0.01) | 1,527 (0.01) |
| | Stroke or TIA | 407,668 (3.56) | 483,295 (3.84) |
| | Thrombosis or pulmonary embolus | 2,458 (0.02) | 3,172 (0.03) |

Data as number (column %) or mean (SD).

* = score from 0 (no exposure) to 100 (maximum exposure).

**eFigure 1. Flow diagram of cohort.**

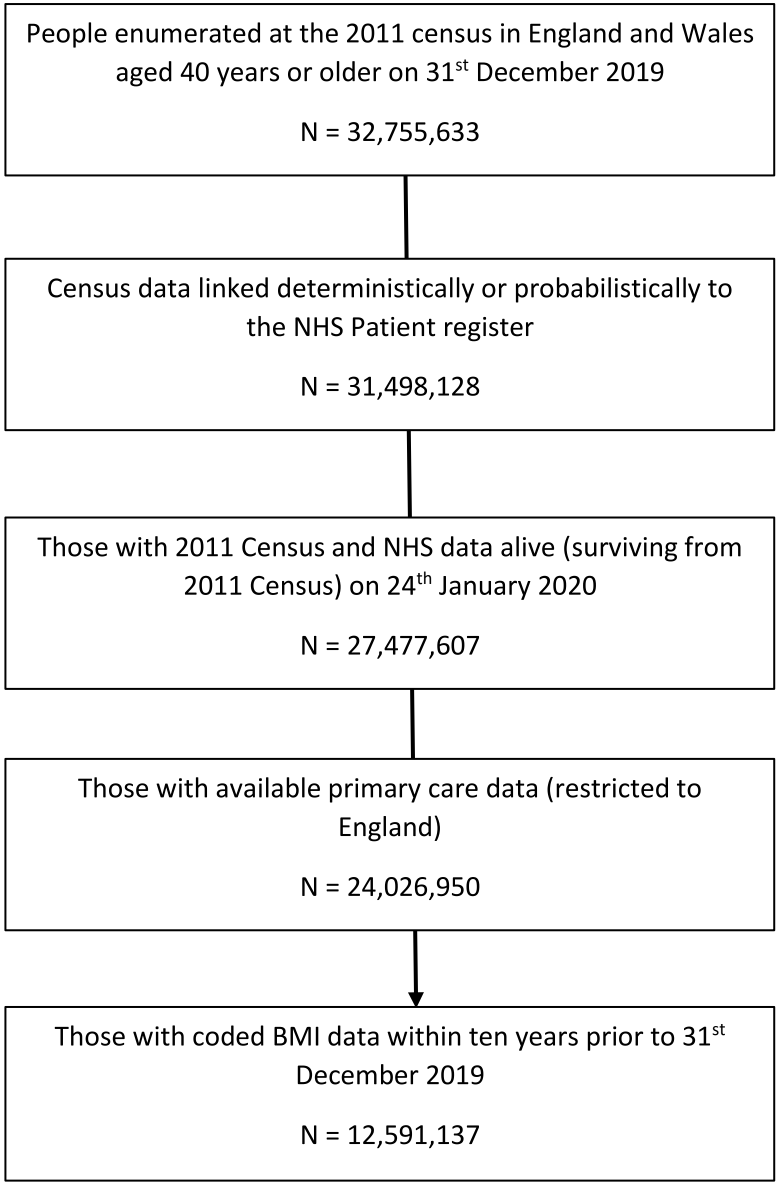

**eFigure 2. Percentage of missing BMI data in each ethnic group by region.**

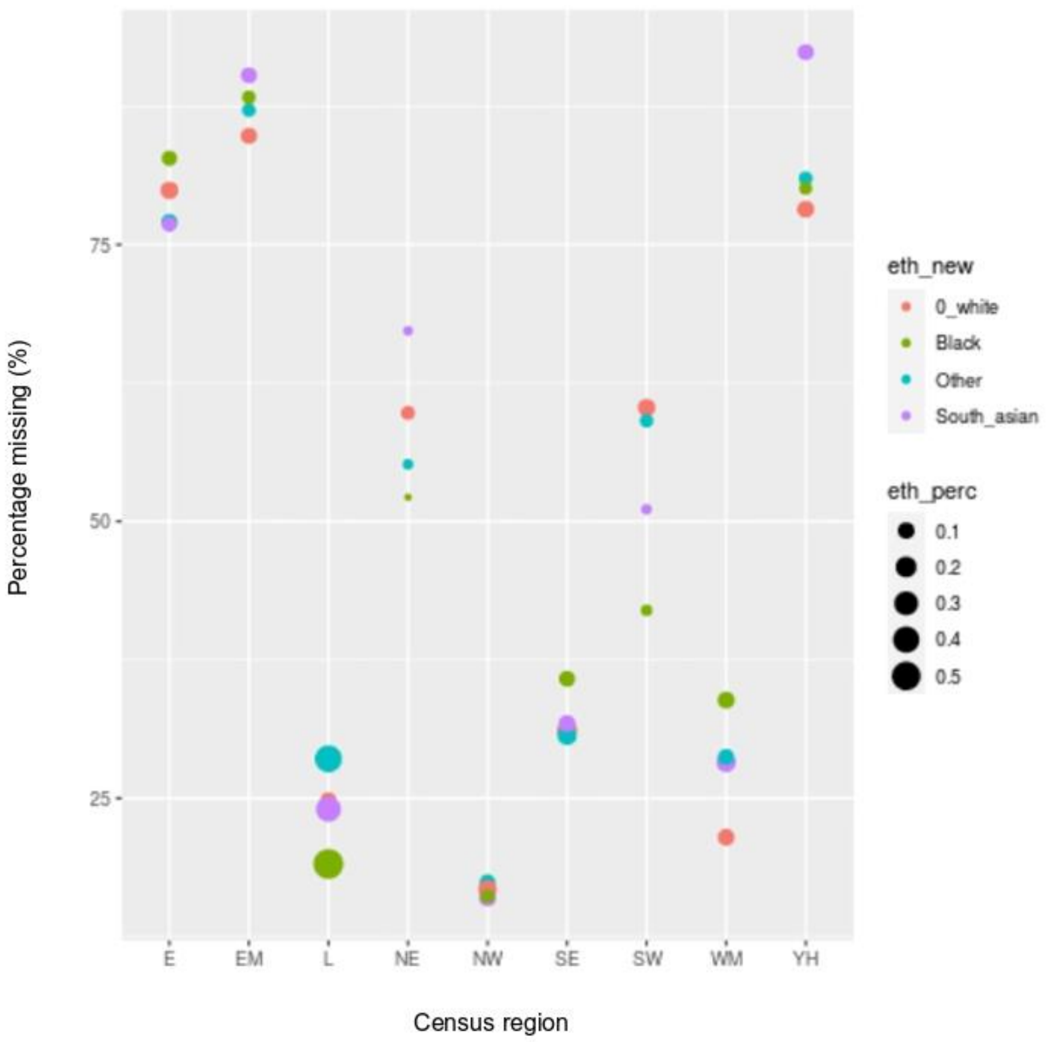

E: East of England; EM: East Midlands; L: London; NE: North East; NW: North West; SE: South East; SW: South West; WM: West Midlands; YH: Yorkshire and Humber.

The size of dot is proportional to the number of people within that specific ethnic group across all nine regions (proportion = 1).

**eFigure 3. Association of BMI with COVID-19 mortality by ethnicity in women.**

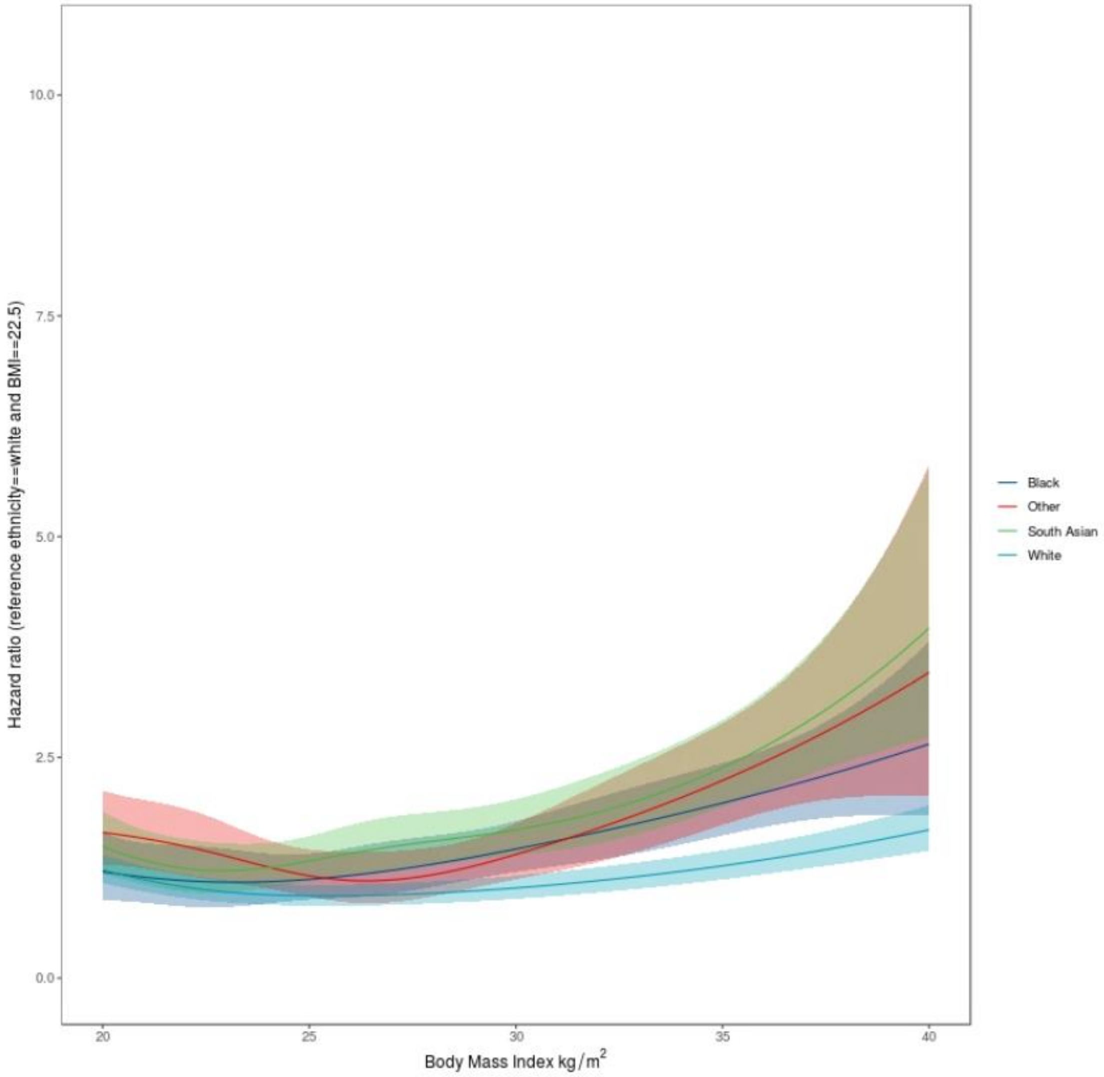

Association of body mass index (BMI) with COVID-19 mortality in white, black, South Asian and 'other' ethnicity women. Hazard ratio (HR) for COVID-19 mortality with BMI stratified by ethnic groups.

Reference (HR 1) were placed at BMI of 22.5 kg/m$^2$ for white individuals. HR (lines) and confidence intervals (CI; areas) are plotted across continuous BMI values (x axes) between the 2.5th (20 kg/m$^2$) and 97.5th (40 kg/m$^2$) centile of the distribution. Shaded area as 95% CI.

Analysis adjusted for: age, sex, region and sociodemographic factors (Model 1 – detailed in Supplementary eTable 1).

**eFigure 4. Association of BMI with COVID-19 mortality by ethnicity in men.**

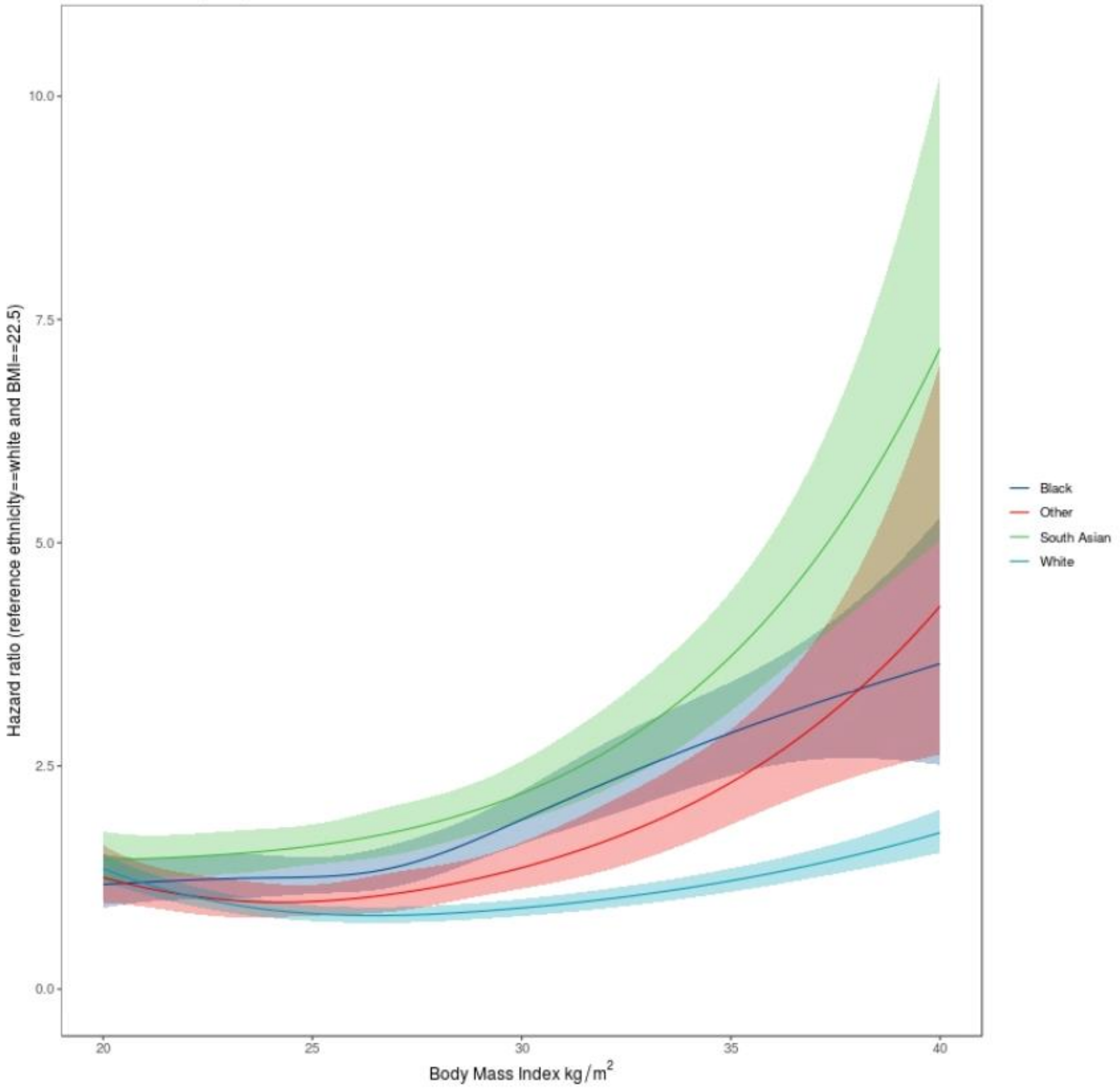

Association of body mass index (BMI) with COVID-19 mortality in white, black, South Asian and 'other' ethnicity men. Hazard ratio (HR) for COVID-19 mortality with BMI stratified by ethnic groups.

Reference (HR 1) were placed at BMI of 22.5 kg/m$^2$ for white individuals. HR (lines) and confidence intervals (CI; areas) are plotted across continuous BMI values (x axes) between the 2.5th (20 kg/m$^2$) and 97.5th (40 kg/m$^2$) centile of the distribution. Shaded area as 95% CI.

Analysis adjusted for: age, sex, region and sociodemographic factors (Model 1 –detailed in Supplementary eTable 1).

**eFigure 5. Association of BMI with COVID-19 mortality by ethnicity: individuals 70 years or greater.**

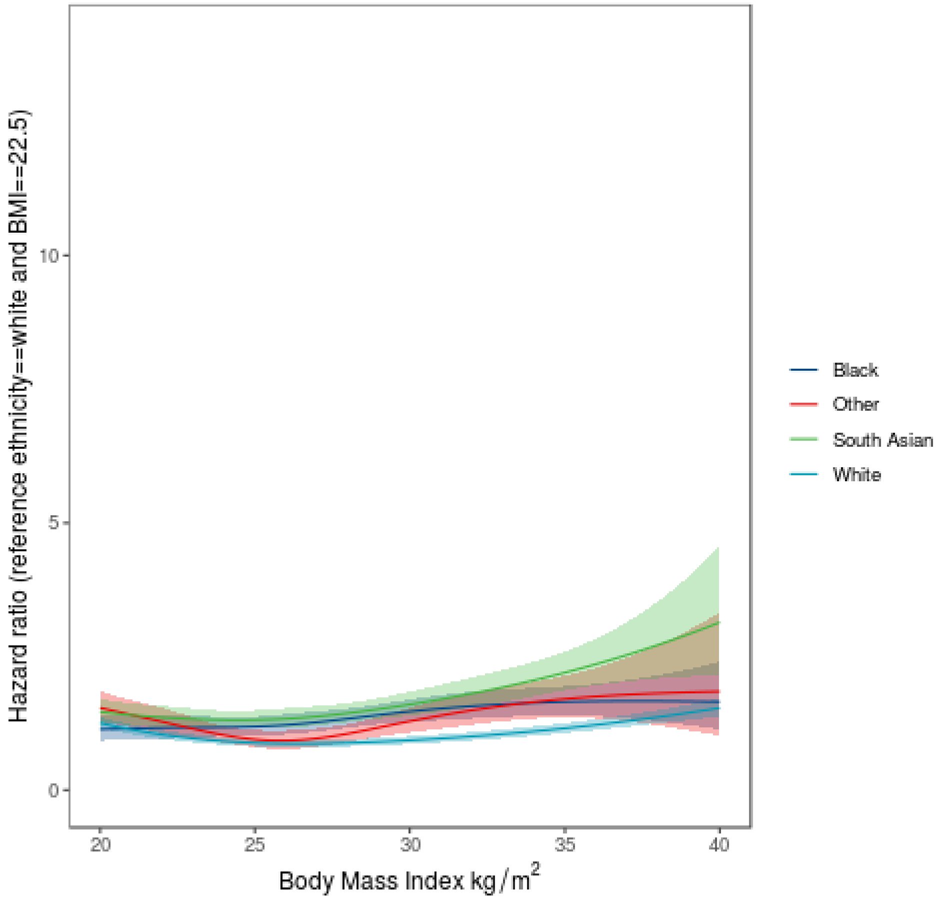

Association of body mass index (BMI) with COVID-19 mortality in white, black, South Asian and 'other' ethnicity individuals. Hazard ratio (HR) for COVID-19 mortality with BMI stratified by ethnic groups.

Reference (HR 1) were placed at BMI of 22.5 kg/m$^2$ for white individuals. HR (lines) and confidence intervals (CI; areas) are plotted across continuous BMI values (x axes) between the 2.5th (20 kg/m$^2$) and 97.5th (40 kg/m$^2$) centile of the distribution. Shaded area as 95% CI.

Analysis adjusted for: age, sex, region and sociodemographic factors (Model 1 –detailed in Supplementary eTable 1).

**eFigure 6. Association of BMI with COVID-19 by ethnicity: individuals younger than 70 years.**

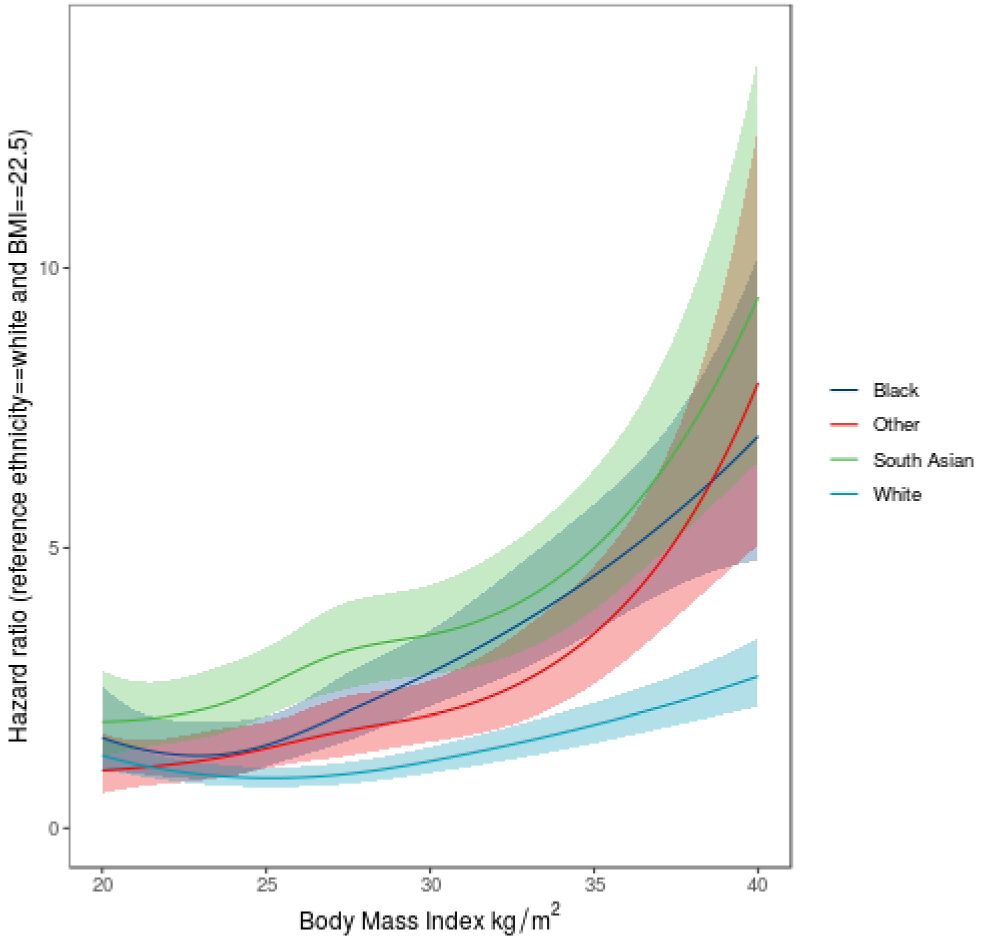

Association of body mass index (BMI) with COVID-19 mortality in white, black, South Asian and 'other' ethnicity individuals. Hazard ratio (HR) for COVID-19 mortality with BMI stratified by ethnic groups.

Reference (HR 1) were placed at BMI of 22.5 kg/m$^2$ for white individuals. HR (lines) and confidence intervals (CI; areas) are plotted across continuous BMI values (x axes) between the 2.5th (20 kg/m$^2$) and 97.5th (40 kg/m$^2$) centile of the distribution. Shaded area as 95% CI.

Analysis adjusted for: age, sex, region and sociodemographic factors (Model 1 – detailed in Supplementary eTable 1)

**eFigure 7. Association of BMI with COVID-19 mortality across ten ethnic categories.**

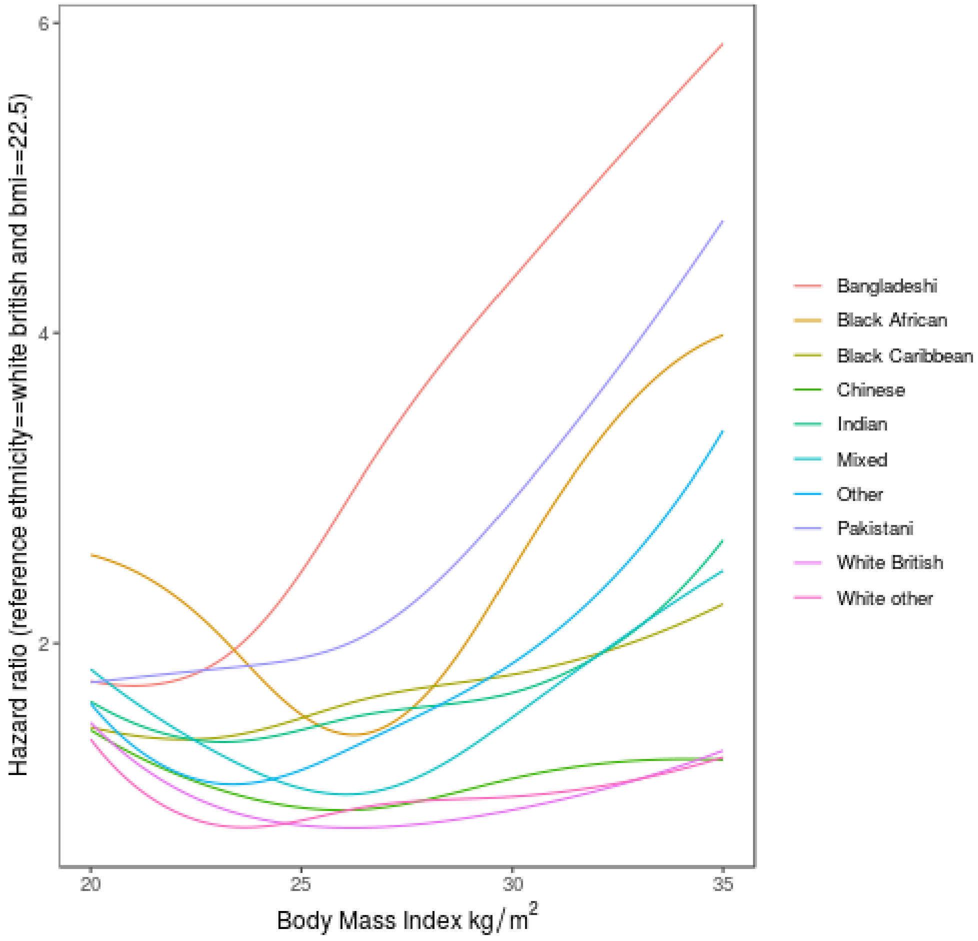

Analysis adjusted for: age, sex, region and sociodemographic factors (Model 1: detailed in Supplementary eTable 1)
